# Cardiac complications following cerebrovascular disease

**DOI:** 10.1101/2023.09.21.23295936

**Authors:** Marco Heinrich Rohner, Catherine Gebhard, Andreas Luft, Martin Hänsel, Susanne Wegener

## Abstract

**Background:** Accumulating evidence suggest that cardiac complications after stroke are an important yet understudied manifestation of brain-heart interactions. Our aim was to investigate and compare cardiac findings after different cerebrovascular events (acute ischemic stroke, transient ischemic attack, and hemorrhagic stroke).

**Methods:** 7113 patients treated between December 2013 and December 2020 at the University Hospital Zurich for ischemic stroke, transient ischemic attack, and hemorrhagic stroke were screened. 721 patients without evidence of previous cardiac disease or presumed cardioembolic origin of their cerebrovascular disease and with at least one cardiac check-up were included. Clinical reports from the year following disease onset were screened for new cardiac findings, which were categorized as arrhythmia/electrocardiographic (ECG) changes, myocardial alterations, valvular abnormalities, coronary perfusion insufficiency, and hypotensive/hypertensive blood pressure abnormalities. Differences in proportions of findings between groups were analyzed using Pearson’s Chi-Square Test or Fisher’s exact Test.

**Results:** ECG changes were observed in 81.7% of patients with ischemic stroke, 71.4% with transient ischemic attack, and 55.8% with hemorrhagic stroke (p<0.001). Myocardial alterations occurred often in all three groups (60.9% ischemic stroke, 59.2% transient ischemic attack, 44.2% hemorrhage, p=0.1). Hypotensive/hypertensive blood pressure abnormalities were most frequent (48.8%) in hemorrhagic stroke patients (p<0.001).

**Conclusions:** Cardiac complications are frequent in patients with cerebrovascular disease, even without prior cardiac problems or suspected cardiac etiology. While findings were different in the three event types, our data suggest that all patients with acute cerebrovascular events should receive thorough workup searching for cardiac complications.

## Introduction

Brain and heart are tightly interconnected. Over the last years, knowledge about the interplay between brain and heart has increased, adding to the growing field of Neurocardiology.^1,2^ On the one hand, cardiac diseases may cause cerebrovascular disease, such as embolization from the heart resulting in ischemic stroke (“heart-brain axis”).^3,4^ On the other hand, findings like cardiac arrhythmia^5^ or damaged myocardial fibers^6^ have frequently been observed to appear after manifestation of acute cerebral lesions (“brain-heart axis”). In fact, increasing evidence suggests that stroke does not only coincide with cardiac pathology due to common risk factors, but instead may provoke cardiac dysfunction.^7–9^ One suggested pathway is the autonomic nervous system impairment, leading to cardiac damage.^9,10^

Different damage mechanisms could lead to disease-specific patterns of cardiac findings after stroke.^9^ However, most studies in this field either focused on a single neurovascular disease entity, ^11,12^ or included patients with preexisting cardiac conditions.^13–15^ Our goal was to determine the fraction and type of cardiac findings, occurring in patients within the year following one of the three types of cerebrovascular disease (CVD): ischemic stroke (IS), transient ischemic attack (TIA) and hemorrhagic stroke (HS). Aiming for cardiac findings after cerebrovascular disease, we excluded patients with known cardiac comorbidities or with suspected cardiac cause of the cerebrovascular event.

## Methods

### Ethics

This study was approved by the local ethics commission of the canton of Zurich (Kantonale Ethikkommission Zürich; KEK-ZH-Nr. 2014-0304). Data from patients were included only if general consent for use of data for research purposes was given.

### Study Settings

This retrospective and descriptive study included stroke patients (IS, TIA or HS) from the Swiss Stroke Registry (SSR) admitted to the Neurology Department at the University Hospital in Zurich (USZ) between the 30^th^ December 2013 and 29^th^ December 2020. “Strengthening the Reporting of Observational Studies in Epidemiology” (STROBE) checklist was used as a reporting guideline^16^.

### Participants

Data from patients in the SSR were screened for three criteria: 1) the presence of TIA, IS or HS, which had to be a first-in-a-lifetime event, 2) records of a neurological check-up at the Department of Neurology within three months after the event, and 3) one cardiological check-up within one year after disease onset were mandatory. We excluded patients with confirmed cardioembolic IS or TIA, also known as “Trial of Org 10172 in Acute Stroke Treatment” subtype two (TOAST 2),^17^ as well as patients with preexisting cardiological or severe neurological diseases (such as severe traumatic brain injury, multiple sclerosis or glioma/brain metastasis). Patients suffering only from minor neurological diseases (e.g., migraine, cognitive impairment, or peripheral polyneuropathy) were still included. A patient flow chart is depicted in Figure 1. In a sub-analysis, we further excluded patients with IS/TIA due to unknown cause (TOAST 5), since this category includes patients with multiple causes of disease, potentially including patients with cardiac source.

**Figure 1.**
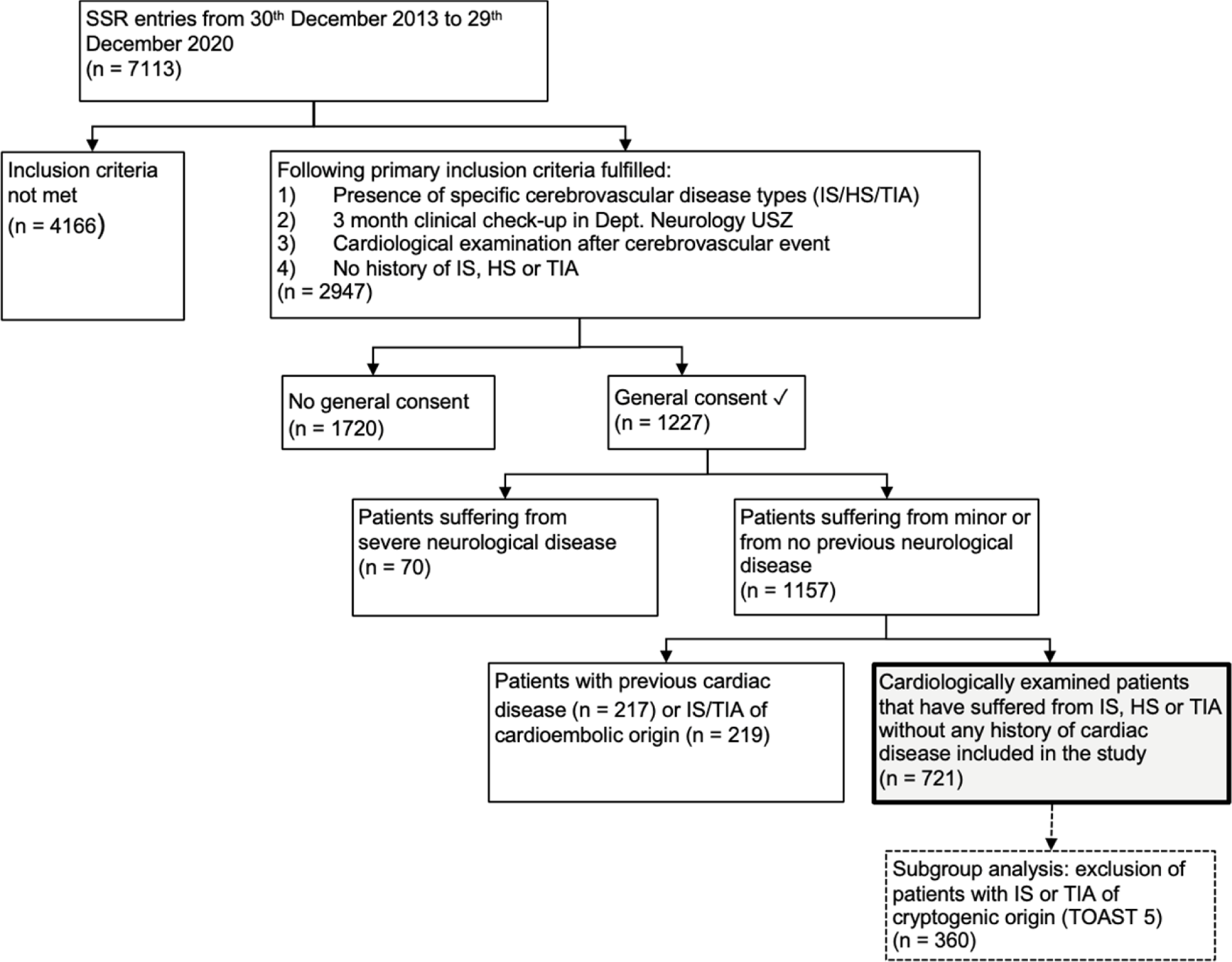
Flow chart showing the exclusion of patients in this investigation. SSR indicates Swiss Stroke Registry; IS, Acute ischemic stroke; HS, Hemorrhagic stroke; TIA, Transient ischemic attack; USZ, University Hospital Zurich; TOAST, Trial of Org 10172 in Acute Stroke Treatment.

### Source of data

Patients fulfilling the criteria mentioned above were eligible for further investigation. All cardiological, neurological, and intensive care reports, created within one year after the cerebrovascular event, were retrieved from the hospital’s clinical information system. Echocardiographic reports as well as Cardiovascular Magnetic Resonance Imaging (CMR), Coronary Computed Tomography Angiography (CCTA) or myocardial perfusion Single Photon Emission Computer Tomography (SPECT) was screened allowing the accurate detection of myocardial findings. Neurological outcome parameters like the NIH-stroke scale (NIHSS) on admission, as well as NIHSS and modified Rankin Scale (mRS) after three months were collected from the SSR.

The data supporting the results of this study can be made available upon reasonable request.

### Cardiac parameters

All medical reports were screened for new cardiac diseases or findings by a medical professional, who had access to all data. Ambulatory cardiology reports, echocardiography, long-term electrocardiography (ECG), and advanced cardiac imaging were scanned for new cardiac findings. To determine whether a finding was truly new, the medical admission report of the hospital stay due to the CVD was verified. A cardiac parameter finding was considered as new, if present in a medical report created within one year after the cerebrovascular event, and never mentioned anywhere previously in the admission letter. The individual findings per subgroups were analyzed and compared. Due to the large number of those individual observations, only cardiac findings observed in ≥5% of patients in at least one CVD subgroup were summarized in tables shown below. Subsequently, all individual cardiac findings retrieved were categorized according to their anatomical and/or functional origin. In total, five cardiac parameter categories were compared, each representing a specific functional entity of the heart: arrhythmogenic findings/ECG changes, myocardial alterations, valve disease, findings of insufficient coronary perfusion, and hypertensive/hypotensive blood pressure abnormalities. Structural (e.g., atrial enlargement) or functional (e.g., heart failure) abnormalities of the myocardium were considered as myocardial alterations. Hypertensive/hypotensive blood pressure abnormalities included findings of altered/pathological venous or arterial blood pressure findings (e.g., pulmonary venous congestions, arterial hypertension, or hypertensive crisis). Findings of insufficient coronary perfusion on the other hand comprised findings ranging from general coronary heart disease to specific findings such as severe stenosis of the left marginal artery.

### Statistical analysis

Descriptive statistics and statistical testing was done using R (R version 4.1.2).^18^ Logical variables are presented as frequencies and percentages, continuous or discrete variables as median and interquartile ranges (IQR). To determine whether the frequencies of logical variables per cerebrovascular subgroups were significantly different, Pearson’s Chi-Square test was used. If the values of ≥20% of all cells in the contingency table of a Chi-Square Test were less than 5, Fisher’s exact Test was applied instead, to maintain statistical accuracy.^19^ When analyzing the differences of continuous or discrete variables in cerebrovascular subgroups (categorical variables), the non-parametric Kruskal-Wallis-Test was applied. P-values ≤ 0.05 were considered statistically significant.

## Results

### Patient characteristics

Of 7113 patients screened, 2947 patients were selected according to the primary inclusion criteria described above (Figure 1). 96.1% of IS patients, 87.2% of TIA patients and 37.2% of HS underwent a cardiological exam. 721 patients met all inclusion criteria. These patients were then divided into three subgroups, according to their CVD (580 IS, 98 TIA and 43 HS).

Patients’ characteristics are displayed in Table 1. TIA patients were significantly older than IS and HS patients (p=0.013). There were more men than women in the IS subgroup (61.2% male patients), while more women suffered a HS (58.2%). Across all three CVD groups, the most common risk factor was hyperlipidemia. The frequencies of risk factors were different: with most smokers in the IS (31.2%), and most dyslipidemia in the TIA group (63.3%) (p=0.023 and p<0.001). HS patients were most severely affected on admission and after three months (median NIHSS on admission: 8 (IQR 11) and NIHSS after three months: 2 (IQR 5) (p<0.001). The median mRS after 3 months was highest, and thus the functional outcome least favorable, in the HS subgroup (2, IQR: 2), followed by the IS (1, IQR 2) and the TIA (0, IQR 0) subgroup (p<0.001). Detailed information about CVD etiology may be found in Table 1.

**Table 1.** Baseline, Clinical, Outcome and Etiology Characteristics. The values shown are medians with interquartile ranges (IQR) or numbers of observations (n) with percentages of patients showing the given finding. NIHSS indicates National Institutes of Health Stroke Scale; mRS, modified Rankin Scale; TOAST, Trial of Org 10172 in Acute Stroke Treatment; NA, not available. For logical variables Pearson’s Chi-Square Test was used to determine whether the proportions were significantly different amongst the three cerebrovascular subgroups. If >20% of the 2×3 contingency table cells show a value <5, Fisher’s exact Test was applied. †The groups were compared using the non-parametric Kruskal-Wallis Test.

| Variable | Acute<br>ischemic<br>stroke<br>(n = 580) | Transient<br>ischemic<br>attack<br>(n = 98) | Hemorrhagic<br>stroke<br>(n = 43) | P-value |
| --- | --- | --- | --- | --- |
| Baseline characteristics: |  |  |  |  |
| Female: n (%) | 225 (38.8) | 44 (44.9) | 28 (58.2) | 0.030 |
| Age: median (IQR) | 64 (20) | 70 (13.5) | 61 (30.5) | 0.013† |
| Clinical characteristics: |  |  |  |  |
| Previous neurological disease: n (%) | 107 (18.4) | 25 (25.5) | 12 (27.9) | 0.110 |
| History of Diabetes mellitus: n (%) | 71 (12.2) | 12 (12.3) | 5 (11.6) | 0.993 |
| History of Hyperlipidemia: n (%) | 329 (56.7) | 62 (63.3) | 10 (23.3) | <0.001 |
| History of Smoking: n (%) | 181 (31.2) | 23 (23.5) | 6 (14.0) | 0.023 |
| Outcome characteristics: |  |  |  |  |
| NIHSS on admission: median (IQR) | 3 (5) | 0 (1) | 8 (11) | <0.001† |
| NIHSS after 3 months: median (IQR) | 0 (2) | 0 (0) | 2 (5) | <0.001† |
| mRS after 3 months: median (IQR) | 1 (2) | 0 (0) | 2 (2) | <0.001† |
| Death after 3 months: n (%) | 10 (1.7) | 0 (0) | 2 (4.7) | 0.135* |
| Etiology of cerebrovascular disease: |  |  |  |  |
| Ischemic stroke & Transient ischemic attack: |  |  |  |  |
| TOAST 1: n (%) | 141 (24.3) | 13 (13.2) | - | p<br><0.001 |
| TOAST 3: n (%) | 82 (14.1) | 8 (8.2) | - |  |
| TOAST 4: n (%) | 68 (11.7) | 5 (5.1) | - |  |
| TOAST 5: n (%) | 289 (49.8) | 72 (73.5) | - |  |
| Hemorrhagic stroke: |  |  |  |  |
| Hypertension: n (%) | - | - | 25 (58.1) | NA |
| Amyloidangiopathy: n (%) | - | - | 2 (4.7) | NA |
| Trauma: n (%) | - | - | 1 (2.3) | NA |
| Vascular lesion: n (%) | - | - | 2 (4.7) | NA |
| Anticoagulant/antithrombotic treatment: n (%) | - | - | 1 (2.3) | NA |
| Unkown: n (%) | - | - | 12 (27.9) | NA |

### Cardiac findings

In total, 138 different individual cardiac findings were found, with all patients showing at least one new finding. Of those 138 findings, only 25 were observed in ≥5% of patients in at least one CVD subgroup. Their frequencies per subgroup are summarized in Table 2.

**Table 2.** Frequencies of those individual cardiac findings that were present in at least 5% of all patients in at least one neurovascular subgroup. Generally, Pearson’s Chi-squared Test was applied for estimating whether the frequencies of the individual cardiac findings in the neurovascular subgroups (IS, HS and TIA) differ significantly from each other. The values are the number of observations (n) with the percentage of patients in the neurovascular subgroup showing the given observation, the p-value or the Odds ratio with its 95% Confidence interval. IS indicates Acute ischemic stroke; TIA, Transient ischemic attack; HS, Hemorrhagic stroke; NA, Not available. *If >20% of the 2×3 contingency table cells show a value <5, Fisher’s exact Test was applied. In all other cases Pearson’s Chi-squared Test was used.

| Variable | Acute<br>ischemic<br>stroke<br>(n = 580) | Transient<br>ischemic<br>attack<br>(n = 98) | Hemor-<br>rhagic<br>stroke<br>(n = 43) | P-value<br>(all 3<br>sub-<br>groups) | P-value<br>(IS vs.<br>TIA) | Odds ratio<br>(IS vs. TIA) |
| --- | --- | --- | --- | --- | --- | --- |
| **Arrhythmogenic findings/**<br>**ECG-changes: n (%)** |  |  |  |  |  |  |
| Supraventricular extrasystole | 393 (67.8) | 57 (58.2) | 7 (16.3) | <0.001 | 0.063 | 1.51 (0.95 to 2.39) |
| Ventricular extrasystole total | 368 (63.4) | 46 (46.9) | 8 (18.6) | <0.001 | 0.002 | 2.06 (1.3 to 3.2) |
| Ventricular extrasystole undetermined | 172 (29.7) | 28 (28.6) | 7 (16.3) | 0.174 | 0.828 | 1.05 (0.64 to 1.76) |
| Atrial tachycardia | 137 (23.6) | 21 (21.4) | 3 (7.0) | 0.040 | 0.635 | 1.13 (0.66 to 2.01) |
| Monomorphic ventricular extrasystole | 84 (14.5) | 8 (8.2) | 0 (0) | 0.008 | 0.091 | 1.9 (0.88 to 4.71) |
| Polymorphic ventricular extrasystole | 77 (13.3) | 6 (6.1) | 1 (2.3) | 0.018 | 0.046 | 2.34 (1 to 6.78) |
| Sinus arrhythmia | 67 (11.6) | 10 (10.2) | 0 (0) | 0.060 | 0.697 | 1.15 (0.56 to 2.6) |
| Supraventricular tachycardia | 62 (10.7) | 11 (11.2) | 1 (2.3) | 0.207 | 0.874 | 0.95 (0.47 to 2.07) |
| Bradycardia | 48 (8.3) | 12 (12.2) | 1 (2.3) | 0.140 | 0.201 | 0.65 (0.32 to 1.39) |
| Ventricular bigeminal rhythm | 34 (5.9) | 4 (4.1) | 0 (0) | *0.215 | *0.637 | 1.46 (0.5 to 5.8) |
| Non-sustained ventricular tachycardia | 26 (4.5) | 9 (9.2) | 0 (0) | 0.042 | 0.052 | 0.46 (0.2 to 1.17) |
| QTc prolongation | 14 (2.4) | 2 (2.0) | 4 (9.3) | *0.026 | *1.0 | 1.19 (0.27 to 10.93) |
| Atrioventricular block (I-III) | 34 (5.9) | 5 (5.1) | 2 (4.7) | 0.913 | 0.765 | 1.16 (0.43 to 3.89) |
| **Myocardial alterations: n (%)** |  |  |  |  |  |  |
| Diastolic heart failure | 153 (26.4) | 23 (23.5) | 10 (23.3) | 0.769 | 0.543 | 1.17 (0.69 to 2.03) |
| Patent foramen ovale | 82 (14.1) | 9 (9.2) | 2 (4.7) | 0.100 | 0.187 | 1.63 (0.78 to 3.82) |
| Left atrial enlargement | 108 (18.6) | 22 (22.4) | 6 (14.0) | 0.467 | 0.373 | 0.79 (0.46 to 1.4) |
| Septal hypertrophy | 60 (10.3) | 8 (8.2) | 0 (0) | 0.073 | 0.506 | 1.3 (0.59 to 3.25) |
| Hypokinesia | 37 (6.4) | 6 (6.1) | 3 (7.0) | 0.982 | 0.923 | 1.04 (0.42 to 3.11) |
| Right atrial enlargement | 23 (4.0) | 7 (7.1) | 1 (2.3) | 0.288 | 0.157 | 0.54 (0.22 to 1.53) |
| Left ventricular heart failure | 27 (4.7) | 2 (2.0) | 3 (7.0) | *0.360 | *0.361 | 2.34 (0.57 to 20.64) |
| Cardiomegaly | 15 (2.6) | 3 (3.1) | 3 (7.0) | *0.254 | *1.0 | 0.84 (0.23 to 4.62) |
| Concentric hypertrophy | 14 (2.4) | 0 (0) | 3 (7.0) | *0.042 | *0.242 | NA |
| **Valve disease findings: n (%)** |  |  |  |  |  |  |
| Mitral valve insufficiency | 64 (11.0) | 13 (13.3) | 2 (4.7) | 0.318 | 0.520 | 0.81 (0.42 to 1.68) |
| Aortic valve insufficiency | 35 (6.0) | 7 (7.1) | 3 (7.0) | 0.897 | 0.674 | 0.84 (0.35 to 2.3) |
| **Hypertensive/hypotensive blood**<br>**pressure abnormalities: n (%)** |  |  |  |  |  |  |
| Arterial hypertension | 63 (10.9) | 11 (11.2) | 10 (23.3) | 0.050 | 0.915 | 0.96 (0.48 to 2.119) |
| Pulmonary arterial hypertension | 16 (2.8) | 2 (2.0) | 3 (7.0) | *0.244 | *0.945 | 1.36 (0.31 to 12.39) |
| Pulmonary congestion | 10 (1.7) | 1 (1.0) | 5 (11.6) | <0.001 | *0.938 | 1.7 (0.24 to 74.59) |

<u>Arrhythmogenic findings/ECG changes</u> were the most common cardiac abnormality in all subgroups: 81.7% of IS patients, 71.4% of TIA patients and 55.8.0% of HS patients developed at least one type of ECG change within one year after CVD onset (p<0.001). These comprised monomorphic and polymorphic ventricular extrasystoles (VES) in 14.5% and 13.3% of IS patients, in 8.2% and 6.1% of TIA patients and in 0% and 2.3% of HS patients, with polymorphic VES finding frequency being significantly different between subgroups (p=0.046). Supraventricular extrasystoles (SVES) were even more common: 67.8% in IS patients, 58.2% in TIA patients and 16.3% in HS patients (p<0.001). Atrial tachycardia frequency differed as well: 23.6% of IS patients, 21.4% of TIA patients and 7% of HS patients experienced atrial tachycardia (p=0.04). A detailed description of ECG finding frequencies are shown in Table 2. The only ECG-change most common in HS patients were QTc-prolongations, found in 9.3% of HS patients, in 2.4% of IS patients and in 2% of TIA patients (p=0.026).

<u>Myocardial alterations</u> occurred in 60.9% of IS, 59.2% of TIA and 44.2% of HS patients (p=0.099, Figure 2). Left atrial enlargement and diastolic dysfunction were very common across all subgroups. Considering wall motion abnormalities, fewer patients were observed to have akinesia than hypokinesia. 7.5% of IS patients, 7.1% of TIA patients and 9.3% of HS patients showed at least one form of wall motion abnormality (p=0.903). The exact frequency of hypokinesia among the three subgroups is illustrated in Table 2.

**Figure 2.**
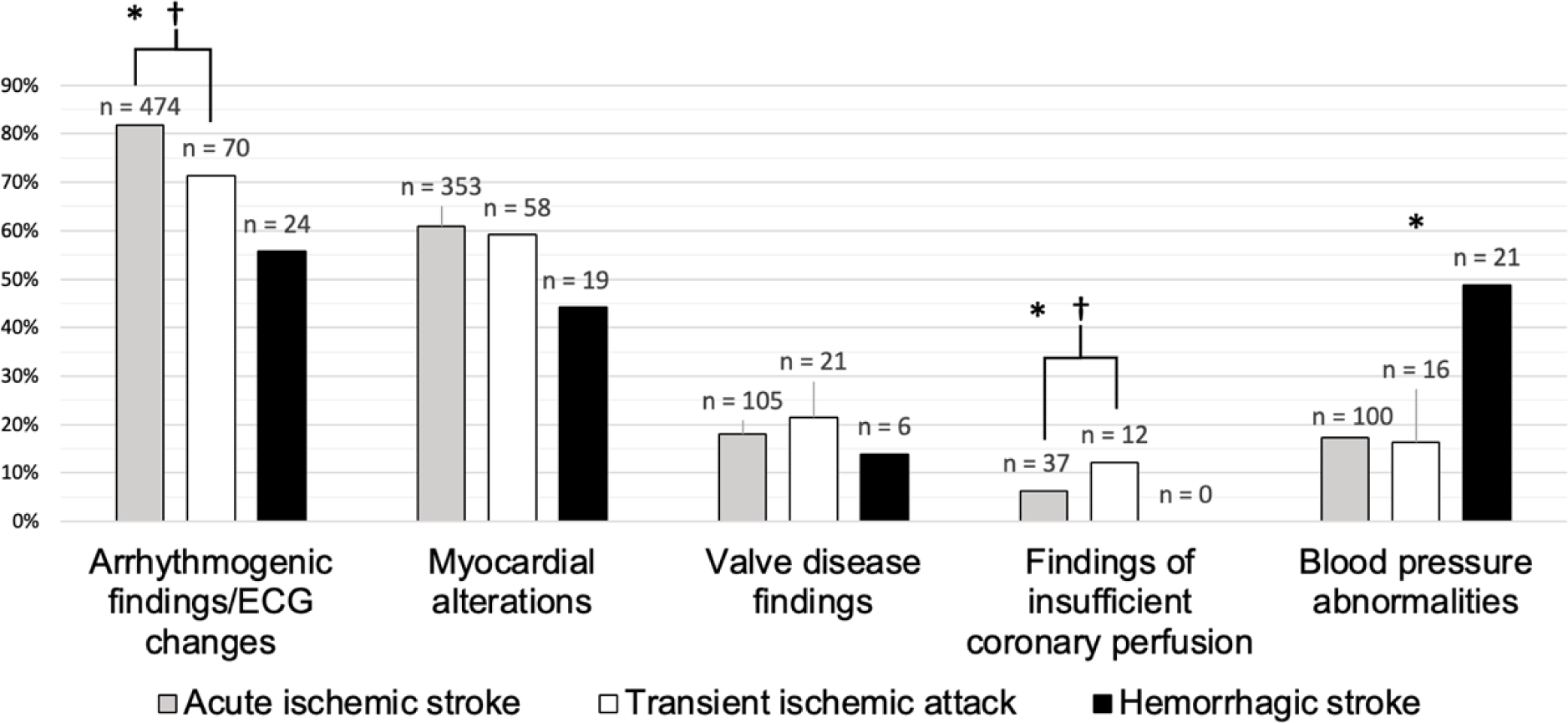
Cardiac findings observed within one year after patients have suffered an IS, HS or TIA, categorized according to their anatomical and functional region of origin. In case the proportions of cardiac findings in the three neurovascular subgroups differ significantly (p ≤ 0.05) from each other (using Pearson’s Chi-Square), the category is marked using an Asterix (*). Further marks (†) are placed, if the proportion of cardiac findings in TIA patients in a given cardiac category differs significantly from the proportion of findings in IS patients.

<u>Valve disease findings</u> were similarly distributed between the three subgroups: 18.1 % of IS, 21.4% of TIA and in 14% of HS patients (p=0.550). The two most common findings were aortic and mitral valve insufficiency (Table 2).

<u>Insufficient coronary perfusion findings</u> were the least prevalent one in all three subgroups. The proportions of findings indicating insufficient coronary perfusion in the three groups were significantly different (6.4% of IS, 12.2% of TIA, and 0% of HS patients, p=0.019). In 2.2 % of IS patients, signs of myocardial infarction within the year following brain ischemia were observed, while they were present in none of the patients from the other two categories. Common individual signs of insufficient coronary perfusion in the IS subgroup were exercise-induced myocardial ischemia (1.2%), ST-segment elevation and depression (1% each), severe coronary sclerosis (1.5%), and myocardial infarct scars (1.6%). TIA patients were diagnosed with exercise-induced ischemia (2%), undetermined coronary sclerosis (3.1%), severe coronary sclerosis (3.1%) and ST-segment depression (3.1%). Altogether 7.1% of TIA patients were diagnosed with some sort of coronary artery sclerosis (by CCTA or interventional coronary angiography), while only 3.6% of IS patients (and none of the HS patients) showed said finding (p=0.099).

<u>New hypertensive/hypotensive blood pressure abnormalities</u> were found in 48.8% of patients suffering from HS. As expected, this was more than in the other two subgroups (17.2% with IS patients and 16.3% with TIA patients (p<0.001)). 10.9% of IS patients, 11.2% of TIA patients and 23.3% of HS stroke patients, were newly diagnosed with arterial hypertension (p=0.050). Pulmonary congestion was most common in HS patients (Table 2, p<0.001).

### Transient ischemic attack vs. acute ischemic stroke

Overall, IS and TIA patients showed very similar frequencies of individual cardiac findings (Table 3). However, when directly comparing IS with TIA, two out of five cardiac parameter categories showed significantly different frequencies: insufficient coronary perfusion findings was more commonly detected in TIA patients (p = 0.038), and arrhythmogenic ECG-changes (p=0.018), more frequently seen in IS patients (Figure 2). VES in general and specifically polymorphic VES were more common in IS. Strikingly, more TIA patients developed coronary stenosis (7.1%), diagnosed by CCTA or interventional coronary angiography, compared to IS patients (3.6%, p=0.178).

**Table 3.**
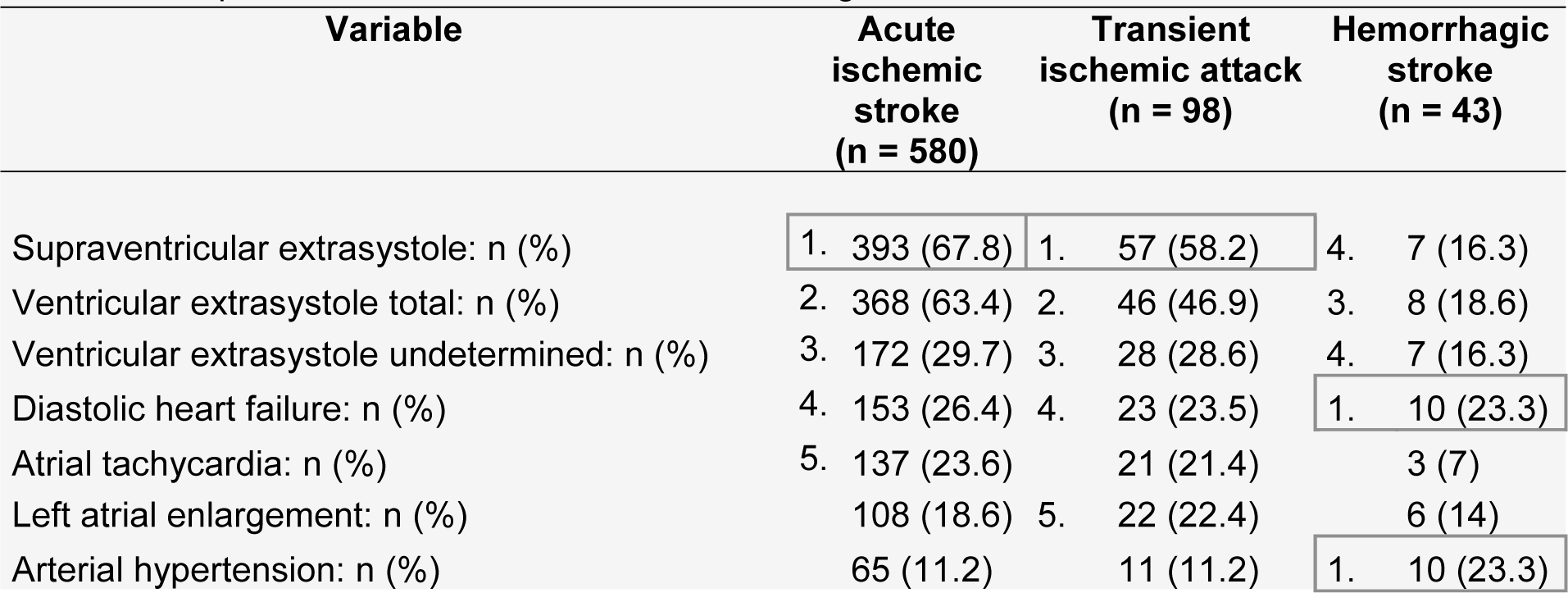
Frequencies of the most common individual cardiac findings per subgroup. This table shows the five most common individual cardiac finding of every subgroup, ranking them according to their proportion of occurrence.

### Subgroup analysis further excluding IS/TIA of cryptogenic origin

49.8% of IS and 73.5% of TIA cases were of cryptogenic origin (Table 1). We did this additional analysis since this group might also include patients suffering from TIA/IS of cardioembolic origin, as TOAST 5 comprises patients with unknown or more than one concurring etiology^17^. Frequencies of ECG-changes remained significantly more common in IS patients (p<0.001), while hypertensive/hypotensive blood pressure abnormalities were still found to be most frequent in HS patients (p<0.001), even when patients with IS and TIA of cryptogenic origin were excluded. However, when directly comparing complications of TIA and IS in this subgroup analysis, significant differences between cardiac parameter groups disappeared. SVES (p<0.001), atrial tachycardia (p=0.034), monomorphic (p=0.009) and polymorphic (p=0.013) VES remained significantly more common in IS and TIA patients than in HS patients.

## Discussion

Arrhythmogenic findings were most frequent, being more common in TIA and IS patients than in HS patients. This observation is particularly important, as cardiac arrhythmia is a frequent cause of death after stroke.^6,9,15^ Our study detected more ECG-changes after stroke than others in IS and TIA patients, which might be explained by the fact that we considered long-term (1 year) follow-up data.^20^ In our investigation, monomorphic and polymorphic VES were most frequent in IS patients, followed by TIA patients. This high prevalence should be kept in mind during the diagnostic workup, as frequent VES, in context of structural heart disease, may increase the probability of sudden cardiac death,^21,22^ and have been associated with increased mortality and higher hospitalization rates, even in patients with structurally normal hearts.^23,24^ Surprisingly, non-sustained ventricular tachycardia was most common in TIA patients (9.2% compared to 4.5% in IS patients). QTc-prolongation was the only ECG-finding most common in HS patients, indicating that this subgroup is at particular risk of developing severe cardiac arrhythmia, such as polymorphic ventricular tachycardia (i.e. torsades de pointes)^25^. Other research groups found similar frequencies of QTc-prolongations in patients with subarachnoid hemorrhage and HS.^26–28^ However, the observation that QTc-prolongations were just as common in TIA patients as in IS patients is new. This and the fact that HS patients seem to be more prone to developing QT-prolongation than IS patients further indicates that long-term cardiac monitoring is warranted in all CVD patients.

The most common myocardial alteration finding was diastolic dysfunction, found in >20% of all patients in every disease subgroup. The average proportion of diastolic dysfunction in the general population aged >65 years was previously estimated between 3.1% and 5.5%.^29^ While left ventricular diastolic dysfunction is known to occur frequently in IS and HS cases,^9,30^ this investigation was able to show a new aspect, with TIA patients showing similarly increased diastolic dysfunction rates. Diastolic dysfunction was proven to be independently associated with functional dependence and death up to 1 year after ischemic stroke^14^ and has therefore to be taken seriously. Left atrial enlargement was observed in ≥14% of all patients in every subgroup. TIA patients were especially prone to developing enlargement of the left atrium (more than every 5^th^ patient). Furthermore, since left atrial enlargement is associated with congestive heart failure^31^, cardiovascular disease^32^, and with recurrent stroke^33,34^, clinicians should incorporate this finding into estimation of prognosis and decisions for secondary prevention. As expected, manifestations of long-standing arterial hypertension (being the cause of >50% of HS patients in this study), cardiomegaly, concentric hypertrophy, left ventricular heart failure and myocardial hypokinesia, were more frequently observed in HS than IS and TIA patients. Cardiomegaly is an independent predictor of death after stroke.^35^ Wall motion abnormalities were observed in >7% of all patients in every subgroup. While the percentage of left ventricular heart failure after TIA matches with previous observations, the finding that non-cardioembolic TIA patients develop wall motion abnormalities to a similar extend as patients with IS may have been underestimated in the past.^36^

HS patients showed the highest frequency of hypertensive/hypotensive blood pressure abnormalities. More than every 5^th^ HS patient showed newly diagnosed arterial hypertension within the year after hemorrhage. Since arterial hypertension is one of the main risk factors for HS^37^, newly diagnosed arterial hypertension might not be a complication, but rather the previously undiagnosed cause of HS. However, patients without any previous chronic hypertension are prone to develop arterial hypertension after HS.^38–40^ Within the IS and TIA subgroup, every 10^th^ patient had newly diagnosed arterial hypertension after CVD. These findings undermine the importance of installing antihypertensive treatments to reduce the risk of reoccurring vascular events after every cerebrovascular disease type.^41^

Compared to the other cardiac categories, signs of insufficient coronary perfusion were not common in TIA and IS patients and virtually inexistent in HS patients. TIA patients showed almost twice as many findings indicating coronary perfusion insufficiency than IS patients. However, this difference disappeared once cryptogenic IS/TIA cases were excluded, which might be a result of the significant reduction of sample size (and therefore statistical power). Insufficient coronary perfusion signs in TIA patients mainly consisted of coronary stenosis. One should bear in mind though that TIA patients were significantly older than the other two subgroups, therefore being more at risk for developing coronary disease.^42^ Our study confirms the high risk of developing myocardial infarction after stroke, with 2.2% of IS patients showing clinical findings indicative of myocardial infarction.^43^ TIA patients merely suffered from ST-segment depression, suggesting myocardial ischemia. Interestingly, no TIA and HS patients showed signs of myocardial infarction. However, this observation must be interpreted with caution, as those two subgroups consisted of significantly fewer patients. Summarizing the above, signs of coronary perfusion insufficiency in previously cardiologically healthy IS patients consisted of myocardial infarction and coronary stenosis. In the TIA subgroup these findings mainly consisted of coronary stenosis, which was, compared to the other subgroups, a very frequent finding.

### Strengths and Limitations

The large number of included patients is a strength of this study. Moreover, the rigorous exclusion of patient suffering from previous diseases or cardioembolic stroke increased the accuracy of new cardiac finding detection. In order to depict cardiac findings as accurately as possible, a large number of medical reports was screened, including those of advanced cardiac imaging, ambulatory cardiac check-ups, echocardiographic examinations, but also reports resulting from a hospital stays in the cardiac unit.

One of the limitations of this study is the different sizes of the three subgroups, rendering statistical comparisons more difficult. Moreover, it should be noted that not every finding newly detected during the cardiological check-up was necessarily new. Additionally, the types of examination performed during cardiological check-ups may vary significantly from patient to patient (as some underwent advanced or even invasive diagnostic steps, such as CCTA or cardiac catheter examinations, while others received only minor diagnostic measures such as long-term electrocardiography). In this investigation, a “new” finding was one that was observed within the year following the cerebrovascular event, without being mentioned in the hospital admission report. While almost all TIA/IS patients received cardiac check-ups, a comparatively small portion of HS patients received said examination. Thus, selection bias (more HS patients in whom cardiac disease might be expected received the workup) is possible in the data from this subgroup. Another minor limitation might be the length of follow-up (12 months), as certain cardiac findings observed might not be stroke-related, but consequences of other independent diseases developed within the year after CVD onset.

## Conclusion

New cardiac findings after non-cardioembolic IS, TIA and HS in patients without preexisting cardiac disease are frequent. IS patients usually have a cardiac follow-up, which this is not the case in many TIA and HS patients (96.1% vs. 87.2% and 37.2%). Findings differ between cerebrovascular disease subgroups. IS patients showed significantly more arrhythmogenic findings/ECG changes than TIA patients, suggesting that structural brain damage might induce these cardiac complications. However, this finding must be interpreted with caution, since after exclusion of all cases with unknown or multiple etiology (TOAST 5), it did not remain significant. Regarding individual cardiac findings, TIA patients and IS patients showed similar results (Table 3). While hypertensive disease in HS patients was expected, ECG changes were frequently observed as well, with QTc-prolongation even being significantly more common than in the other CVD subgroups. Therefore, cardiac workup of patients not only with IS, but also with HS and TIA, is essential to prevent cardiac complications or even death in these patients.

## Data Availability

The data supporting the results of this study can be made available upon reasonable request.

## Acknowledgements

None.

## Non-standard Abbreviations and Acronyms

CVD: Cerebrovascular disease
IS: Acute ischemic stroke
TIA: Transient ischemic attack
HS: Hemorrhagic stroke
SSR: Swiss Stroke Registry
USZ: University Hospital of Zurich
TOAST: Trial of Org 10172 in Acute Stroke Treatment
CCTA: Coronary Computed Tomography Angiography
CMR: Cardiovascular Magnetic Resonance Imaging
SPECT: Single Photon Emission Computer Tomography
NIHSS: NIH-stroke scale
mRS: Modified Rankin Scale
ECG: Electrocardiography
IQR: Interquartile range
VES: Ventricular extrasystole
SVES: Supraventricular extrasystole

